# No evidence for differential saccadic adaptation in children and adults with an Autism Spectrum diagnosis

**DOI:** 10.1101/2023.05.31.23290682

**Authors:** Katy Tarrit, Edward G. Freedman, Ana Alves Francisco, Douwe J. Horsthuis, Sophie Molholm, John J. Foxe

## Abstract

**Background:** Altered patterns of eye-movements during scene exploration, and atypical gaze preferences in social settings, have long been noted as features of the Autism phenotype. While these are typically attributed to differences in social engagement and interests (e.g., preferences for inanimate objects over face stimuli), there are also reports of differential saccade measures to non-social stimuli, raising the possibility that fundamental differences in visuo-sensorimotor processing may be at play. Here, we tested the plasticity of the eye-movement system using a classic saccade-adaptation paradigm to assess whether individuals with ASD make typical adjustments to their eye-movements in response to experimentally introduced errors. Saccade adaptation can be measured in infants as young as 10 months, raising the possibility that such measures could be useful as early neuromarkers of ASD risk.

**Methods:** Saccade amplitudes were measured while children and adults with ASD (N=41) and age-matched typically developing (TD) individuals (N=68) made rapid eye-movements to peripherally presented targets. During adaptation trials, the target was relocated from 20-degrees to 15-degrees from fixation once a saccade to the original target location was initiated, a manipulation that leads to systematic reduction in saccade amplitudes in typical observers.

**Results:** Neither children nor adults with ASD showed any differences relative to TD peers in their abilities to appropriately adapt saccades in the face of persistently introduced errors.

**Conclusions:** Of the three studies to date of saccade adaptation in ASD, none have shown frank deficits in saccade adaptation. Unlike prior studies, we found no evidence for a slower adaptation rate during the early adaptation phase, and no of evidence greater variance of saccade amplitudes in ASD. In post-hoc analysis, there was evidence for larger primary saccades to non-adapted targets, a finding requiring replication in future work.

## INTRODUCTION

While Autism is primarily associated with atypical social communication and restricted/repetitive behaviors, sensory-motor differences are also very commonly reported (Brenner, Turner, & Müller, 2007; Fournier, Hass, Naik, Lodha, & Cauraugh, 2010; Molloy, Dietrich, & Bhattacharya, 2003; Nowinski, Minshew, Luna, Takarae, & Sweeney, 2005; Y. Takarae, Minshew, Luna, & Sweeney, 2004). For example, postural instability, coordination difficulties, and atypical oculomotor control have all been shown in this population (Brenner et al., 2007; Ghaziuddin & Butler, 1998; Jones & Prior, 1985; C. Kemner, Verbaten, Cuperus, Camfferman, & Van Engeland, 1998; Klin, Jones, Schultz, Volkmar, & Cohen, 2002; Kohen-Raz, Volkman, & Cohen, 1992; Molloy et al., 2003; Nowinski et al., 2005; Rogers, Bennetto, McEvoy, & Pennington, 1996; Stanley-Cary, Rinehart, Tonge, White, & Fielding, 2011; Y. Takarae et al., 2004; Vilensky, Damasio, & Maurer, 1981). Oculomotor functions are of particular interest given that accurate eye movements govern integration between the visual and other sensory systems, are often essential for motor planning, and are implicated in a multitude of other processes potentially relevant to the Autism phenotype, such as visual search, imitation, joint attention, and language (Brenner et al., 2007).

In the context of Autism, eye-movements have been of substantial interest to clinicians and researchers because of the long-noted atypicalities that autistic individuals show in making or maintaining eye-contact in social settings (Hirsch et al., 2022). A key question arises regarding the nature of these gaze differences – that is, 1) are they driven by higher-order social communication difficulties such that they reflect a reluctance to engage socially, or 2) is there a more fundamental oculomotor processing deficit that underlies these differences. Here, we were interested in testing this second thesis that more basic oculomotor physiological differences may exist in autism spectrum disorder (ASD), independent of engagement with socially relevant inputs. We have previously hypothesized that differences in visuo-sensorimotor development in early infancy may underlie some of the differences in visual orienting, communication and social interaction reported in Autism, perhaps as a result of structural cerebellar differences that impact plasticity of the saccadic eye-movement system (Freedman & Foxe, 2018). We set out to test whether simple saccadic eye-movements to basic non-social stimuli, in the context of a saccade adaptation paradigm, would differ in individuals with an Autism diagnosis relative to matched typically developing peers.

Saccades are rapid conjugate eye movements that shift the direction of gaze from one location to another in order to accurately aim the highly-innervated fovea at areas or objects of interest (Fuchs, Kaneko, & Scudder, 1985). Modern infrared camera techniques allow for accurate measurement of these eye-movements, and their dynamics and neural substrates have been studied extensively (N Alahyane et al., 2008; Belyusar et al., 2013; Collins & Wallman, 2012; J. J. Hopp & Fuchs, 2004; Kelly, Foxe, Newman, & Edelman, 2010). Once a visual target has been identified, its position in relation to the fovea is calculated and sent as a motor command to activate the extraocular muscles so that the eyes move in the correct direction and to the appropriate location to fixate said target. At the end of a saccadic eye movement, if the eye-movement displacement is erroneous, either undershooting or overshooting the target location, the vector of a subsequent saccade is calculated and a new motor command generated by the oculomotor system to adjust for the error. It is this oculomotor learning process that has been termed saccade adaptation (J. J. Hopp & Fuchs, 2004; Pélisson, Alahyane, Panouilleres, & Tilikete, 2010), allowing the oculomotor system to adjust and maintain the amplitude and direction of saccades by adaptatively adjusting motor commands in response to persistent visual errors (Lappe, 2009; Wong & Shelhamer, 2011).

Saccade adaptation is often produced in the laboratory by using paradigms in which the target is surreptitiously moved while the eye is in flight, such as in the McLaughlin paradigm (McLaughlin, 1967) and its variations (Nadia Alahyane & Pélisson, 2004, 2005; Cecala & Freedman, 2008, 2009; Deubel, Wolf, & Hauske, 1986; Frens & Van Opstal, 1997; J Johanna Hopp & Fuchs, 2002, 2006; Miller, Anstis, & Templeton, 1981; Noto, Watanabe, & Fuchs, 1999; Phillips, Fuchs, Ling, Iwamoto, & Votaw, 1997; Robinson, Noto, & Bevans, 2003; Scudder, Batourina, & Tunder, 1998; Straube, Fuchs, Usher, & Robinson, 1997; Takeichi, Kaneko, & Fuchs, 2005; Takeichi, Kaneko, & Fuchs, 2007; Wallman & Fuchs, 1998). It is well-established that the midline cerebellum plays a critical role in saccade adaptation (Avila et al., 2015; Barash et al., 1999; Galea, Jayaram, Ajagbe, & Celnik, 2009; Gaymard, RivaudlJPechoux, Yelnik, Pidoux, & Ploner, 2001; Jayaram et al., 2012; Panouillères, Miall, & Jenkinson, 2015; Straube, Deubel, Ditterich, & Eggert, 2001). In non-human primates, studies have demonstrated that lesioning of the oculomotor cerebellar vermis causes acute hypometria of saccades as well as a severe and relatively chronic deficit in saccade adaptation (Barash et al., 1999) and that inactivation of the caudal fastigial nucleus through acute muscimol injections leads to aberrant saccade dynamics and inaccurate fixations (Goffart, Chen, & Sparks, 2004). In humans, cerebellar degeneration, infarcts or congenital malformations also lead to disruption of saccade amplitude (Straube et al., 2001).

This is relevant to ASD because there is a large literature reporting potential neuroanatomical differences in the structure of the cerebellum in individuals with ASD (Courchesne et al., 1994; Courchesne, Yeung-Courchesne, Hesselink, & Jernigan, 1988; Nowinski et al., 2005; Stanley-Cary et al., 2011; Y. Takarae et al., 2004). One study using magnetic resonance imaging found that individuals with ASD have significantly smaller neocerebellar lobules VI and VII compared to their typically developing (TD) peers (Courchesne et al., 1988). Another study reported that there were two subgroups in the ASD population, one with vermian hypoplasia and one with vermian hyperplasia (Courchesne et al., 1994). As the cerebellum contributes significantly to visuomotor processing, it is a reasonable proposition that saccadic eye movements might be affected by these structural differences, which would go to our primary thesis here that eye-movement atypicalities in ASD may be due to basic sensory-oculomotor integration differences rather than to a failure to engage in socially relevant interactions.

There is a substantial emerging literature investigating oculomotor functions in ASD by measuring saccade dynamics and metrics, but these have yielded mixed and sometimes contradictory results (Nico Bast et al., 2021; Glazebrook, Gonzalez, Hansen, & Elliott, 2009; Goldberg et al., 2002; B. Johnson et al., 2012; Chantal Kemner, Van der Geest, Verbaten, & van Engeland, 2004; C. Kemner et al., 1998; Kovarski, Siwiaszczyk, Malvy, Batty, & Latinus, 2019; Luna, Doll, Hegedus, Minshew, & Sweeney, 2007; Minshew, Luna, & Sweeney, 1999; M. Mosconi et al., 2009; Pensiero, Fabbro, Michieletto, Accardo, & Brambilla, 2009; Rosenhall, Johansson, & Gillberg, 1988; Schmitt, Cook, Sweeney, & Mosconi, 2014; Stanley-Cary et al., 2011; Yukari Takarae, Luna, Minshew, & Sweeney, 2008; Y. Takarae et al., 2004; Thakkar et al., 2008; van der Geest, Kemner, Camfferman, Verbaten, & van Engeland, 2001; Wilkes, Carson, Patel, Lewis, & White, 2015). While some have reported atypical saccade dynamics, others find no obvious differences between ASD and TD populations. Some have reported longer saccade latencies (Glazebrook et al., 2009; Goldberg et al., 2002; Pensiero et al., 2009; Schmitt et al., 2014; Wilkes et al., 2015), while others found no significant differences (B. P. Johnson, Lum, Rinehart, & Fielding, 2016; Kovarski et al., 2019; Minshew et al., 1999). Some studies found a decrease in saccade duration (N. Bast et al., 2021), and others found significantly longer saccade durations (Stanley-Cary et al., 2011). Other studies have also reported a decrease in saccade amplitude, i.e., hypometria, (N. Bast et al., 2021; B. Johnson et al., 2012; Luna et al., 2007; Rosenhall et al., 1988; Y. Takarae et al., 2004) while some found no differences in mean accuracy (B. P. Johnson et al., 2016). Similarly, some reported more variability in saccade amplitude or latency (Glazebrook et al., 2009; Goldberg et al., 2002; B. Johnson et al., 2012; Stanley-Cary et al., 2011; Y. Takarae et al., 2004) or an increase in saccadic activity (C. Kemner et al., 1998). Others found reduced (Rosenhall et al., 1988) or irregular (Pensiero et al., 2009) saccade peak velocity but similar reaction times for individuals with ASD compared to their TD peers (Rosenhall et al., 1988), while some studies reported no group differences in peak velocity (B. P. Johnson et al., 2016; Luna et al., 2007). In one study, ASD children were found to be as accurate but faster than TD children in looking toward targets (Kovarski et al., 2019). Studies using other tasks such as the anti-saccade task have also reported impaired performance in ASD (Goldberg et al., 2002; M. Mosconi et al., 2009; Thakkar et al., 2008).

To our knowledge, only two studies have investigated saccade adaptation in ASD to date (B. P. Johnson, Rinehart, White, Millist, & Fielding, 2013; M. W. Mosconi et al., 2013). Johnson and colleagues conducted a study on children between 9-14 years of age and found that both individuals with High Functioning Autism (HFA) and those classified as Asperger’s (AD) adapted appropriately compared to a TD control group. However, they also reported that the HFA group showed a slower adaptation rate (B. P. Johnson et al., 2013), although it is important to point out that this latter conclusion was reached on the basis of a comparison between cohorts of 10 individuals with HFA and 12 TD individuals. In a much larger study across a considerably larger age-range (8-54 years), however, Mosconi and colleagues also reported that their ASD group (N=56) adapted slower than the TD group (N=53). In addition, they reported that almost a third of their ASD cohort did not adapt at all, versus only 6% of their TD group. Their ASD group also showed more variability in saccade amplitudes during each trial type, i.e. baseline, adaptation and recovery (M. W. Mosconi et al., 2013). Given the paucity of existing data on saccade adaptation in ASD and the lack of entirely consistent results across the two existing studies, we set out here to investigate whether saccade adaptation would be atypical in both children (N=21: 6-17 years of age) and adults (N=20: 18-45 years of age) on the Autism Spectrum compared to their TD peers in the context of the so-called “gain-down” saccade adaptation paradigm, a variant of the one pioneered by McLaughlin (McLaughlin, 1967).

A main motivation for this work is that it has been shown that saccade adaptation can be measured in infants as young as 10 months of age (Lemoine-Lardennois et al., 2016). As such, if saccade adaptation deficits are consistently identified in ASD, this relatively simple-to-deploy measure could prove useful as part of an early risk-assessment battery (Goldberg et al., 2002; Wilkes et al., 2015). We were also specifically interested in assessing whether saccade adaptation deficits might be more severe in children than adults with ASD. While Mosconi and colleagues found no association between age and saccade adaptation parameters in their study, we specifically recruited separate groups of children and adults to directly address this issue, since developmental effects across the adult lifespan may have contributed to the relative lack of saccade adaptation deficits that have been seen in the prior work.

## METHODS

### Participants and Phenotypic Assessments

41 individuals with an Autism Spectrum Diagnosis (ASD) and 68 age-matched Typically Developing (TD) controls (age range: 6 to 45 years old) were recruited (see Table 1 for demographics). Participants all had normal or corrected-to-normal vision (20/30). Participants with corrected vision were asked to remove their glasses and wear COMO SPORT L glasses with lenses adjusted to their visual acuity for the duration of the experiment. Participants were excluded from the study if they had a history of seizures, traumatic brain injury, psychiatric or neurodevelopmental conditions other than autism (not including ADHD or anxiety), or if they were taking any antiepileptic or anticonvulsant medication, as these can impact eye movements (Aschoff, 1968; Cohen et al., 1985; Goldman & Schultz-Ross, 1993; Hilton, Hosking, & Betts, 2004; Lo, Shorvon, Luxon, & Bamiou, 2008; Lunn et al., 2016; Park et al., 2015; Reilly, Lencer, Bishop, Keedy, & Sweeney, 2008; Remler, Leigh, Osorio, & Tomsak, 1990; Tedeschi et al., 1989; Zaccara et al., 1992). The current study was approved by the Institutional Review Boards (IRB) of the University of Rochester Medical Center and of the Albert Einstein College of Medicine. Written informed consent was obtained from participants or their parent/legal guardian. Assent appropriate for age and developmental level was obtained from participants where applicable. Participants were compensated at a rate of $15 per hour for their time in the laboratory.

**Table 1.**
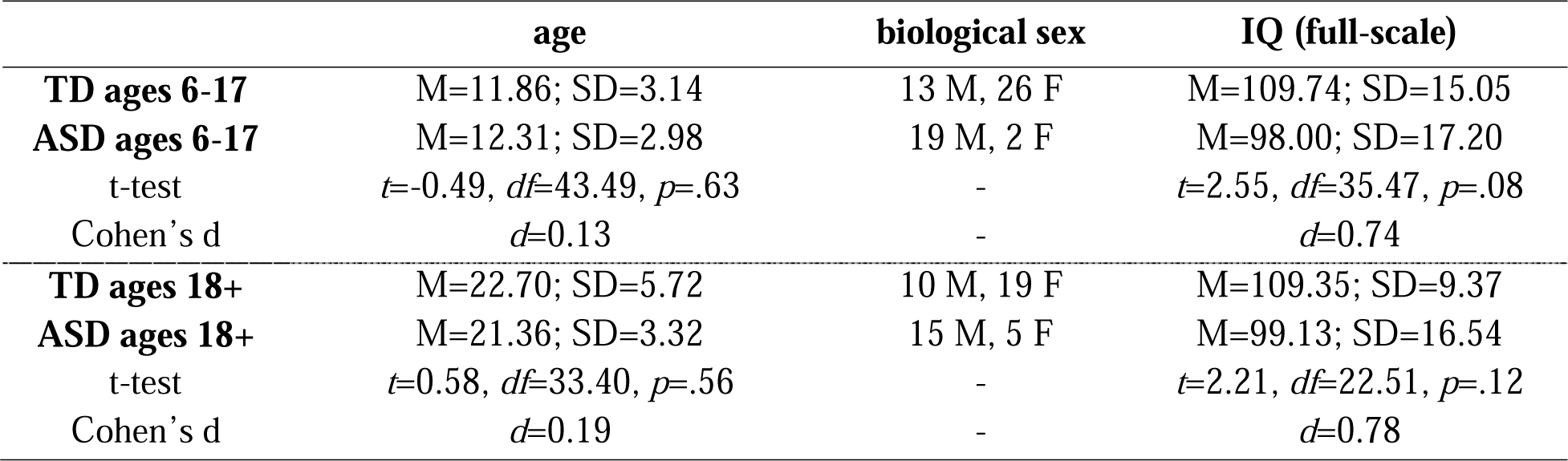
Characterization of the typically developing and ASD groups included in the analyses: Age, biological sex, and IQ.

Phenotyping sessions lasted about one and a half hours for TD participants and two hours for individuals with ASD. The Wechsler Abbreviated Intelligence Scale, 2nd Edition (WASI-II) (Wechsler, 2011) was used to assess verbal and non-verbal abilities. Those reporting a diagnosis of ASD additionally completed Module 3 or Module 4 of the Autism Diagnostic Observation Schedule, 2nd Edition (ADOS-2) (Gotham, Risi, Pickles, & Lord, 2007). Individuals with ASD were assessed by a clinical psychologist, research reliable on the ADOS-2. TD participants were assessed by either a clinical psychologist or a post-doctoral fellow supervised by a clinical psychologist. Two participants who were initially recruited into the autism group were excluded from the analysis, as despite valid autism diagnosis by a qualified professional based on DSM criteria at a younger age, these individuals no longer met instrument classification for ASD on the ADOS-2. All individuals in the ASD group had a positive ADOS-2 score and met DSM-5 criteria for autism.

### Stimuli and Experimental Task

Eye movements were recorded using the video-based eye tracker EyeLink 1000 (SR Research, Ottawa, Canada), with an average accuracy of 0.15 degrees and a sampling rate of 1000 Hz. Only one eye was monitored. The amplitudes of primary saccades (i.e., the initial eye movements from the centrally presented fixation cross (T0) to the target location (T1)) were identified and extracted off-line using a custom MATLAB script (MathWorks, Natick, MA). Noisy trials were excluded and only trials without blinks and in which participants looked toward the intended target were included in the analysis. The amplitude of primary saccades was then plotted as a function of the trial number, i.e., the order in which they occurred during the experiment, including control trials—which occurred pre- and post-adaptation trials.

During the experiment, participants sat on a chair facing the center of a computer monitor, placed 78.8 cm away from the participant, where stimuli were presented. At the beginning of the experiment, the camera was calibrated on the selected eye by asking the participant to make saccades to nine fixation crosses randomly presented either at the center of the screen, in each direction along the middle line (top, bottom, left and right of the center) or along the corners. The experimental paradigm was a gain-down saccadic adaptation paradigm adapted from the one introduced by McLaughlin (McLaughlin, 1967) and consisted of the presentation of a series of 68 pre-adaptation or control trials, followed by a series of 240 adaptation trials, interspersed with control trials, and a final series of 68 post-adaptation or control trials.

Initial control trials consisted of the presentation of a central target (T0) that participants were required to fixate within ±0.75° for a variable interval of either 600, 800, 1000 or 1200 ms. Breaking fixation resulted in aborting the trial and the beginning of a new trial. At the end of this interval, the T0 target was turned off and a secondary target (T1) was presented at one of the following locations: either 20° to the left or to the right of the T0 location, or 10° above or below T0. Participants were then instructed to saccade as accurately as possible to the T1 target. As soon as they started a saccade towards T1, the T1 target disappeared and after a fixed interval of 1500 ms the next trial started. After 68 of these control trials, the series of adaptation and interspersed control trials started. Adaptation trials were similar to control trials. The difference was that adaptation trials were only presented on the right side of the central target, and after T1 was turned off, it was relocated to a T2 target located 15° from the central target, i.e. 5° closer to T0 compared to T1. The T2 target was then displayed for an interval of 500 ms before it was turned off followed by a blank screen for an interval of 1000 ms before the start of the next trial. During the series of adaptation trials, control trials in other directions related to the center of the screen (left, top or bottom) were randomly interspersed to prevent participants from predicting the location of the next target. After presentation of 200 adaptation trials, a set of 68 post-adaptation control trials were presented. This involved presentation of the T1 at 20 degrees without any subsequent movement, the same as the original pre-adaptation control trials. This final series of control trials served as a recovery phase.

### Analytical approach

Multiple saccade metrics were of interest: 1) the amplitude of primary saccades, 2) the early and the late phase of adaptation (as defined below), 3) the mean amplitude of all control trials which occurred to the left of the central target during the entire experiment (i.e., during both series of control trials and the series of adaptation trials mixed with control trials), and 4) the within-participant variability in saccade amplitude to initial control trials. The early phase was calculated as the difference between the mean amplitude of the last twenty pre-adaptation control trials and the mean amplitude of the first twenty adaptation trials, while the late phase was calculated as the difference between the mean amplitude of the last twenty pre-adaptation control trials and the mean amplitude of the last twenty adaptation trials. A two-way repeated measures ANCOVA was performed to test for differences in adaptation between the two groups as well as to investigate the impact of age on saccadic adaptation across groups. A univariate ANCOVA was used to determine if eye movement amplitudes differed between individuals with ASD and their age-matched controls, i.e., to verify if individuals with ASD tended to undershoot (hypometria) or overshoot (hypermetria) the visual target. Finally, an independent samples t-test was performed to look at the variability of eye movements in each group. Variability was calculated by deriving the standard deviation of the last twenty pre-adaptation control trials for each individual separately, and normalized by dividing this by the individuals’ mean saccade amplitude across those last twenty trials. All analyses were accomplished using MATLAB (MathWorks, Natick, MA) and SPSS (IBM, Chicago, IL).

## RESULTS

For illustrative purposes, Figure 1 provides examples of adaptation from two TD participants and two individuals with an ASD diagnosis, providing an example of a faster and a slower adapter for each group. Trials to the left (in green) correspond to the first series of control trials (the baseline phase), trials in the middle (in yellow) are adaptation trials during which the systematic shift of the target was introduced, and trials to the right (in green) correspond to the final series of control trials (i.e., the recovery phase), which was identical to the initial baseline phase. In the case of the faster TD adapter (TD1, top-right panel), the participant made saccades to the T1 target with a mean amplitude of 17.56 (SD=0.72) during the baseline phase, but within the first 20 adaptation trials, the mean amplitude decreased to 16.19 (SD=0.83) as a result of the introduction of the inward target shifts. This decrease in saccade amplitude continued to develop over the remaining adaptation trials, until the mean amplitude of the last 20 adaptation trials reached 11.83 (SD=2.98). For the slower adapting TD (TD2, bottom-right panel), saccades to the target during baseline had a mean amplitude of 18.93 (SD=0.93). During the first 20 adaptation trials, the mean amplitude remained high at 19.52 (SD=0.87), showing no evidence for adaptation to the introduced errors. Over the remaining adaptation trials, the mean amplitude decreased gradually to 16.57 (SD=0.91). Likewise, for the sample rapid-adapter from the ASD cohort (ASD1, top-left panel), a mean saccade amplitude of 16.38 (SD=1.12) was observed during baseline trials. Within the first 20 adaptation trials, the mean amplitude decreased to 14.68 (SD=1.03) and this decrease continued to evolve over subsequent adaptation trials until the mean amplitude reached 13.62 (SD=0.77). In the case of the slow ASD adapter (ASD2, bottom-left panel), baseline trials had a mean amplitude of 15.93 (SD=1.34), with little change seen during the initial 20 adaptation trials (Mean = 15.55; SD=1.39), and only a very modest decrease observed over the subsequent adaptation trials (Mean = 13.78; SD=1.30).

**Figure 1.**
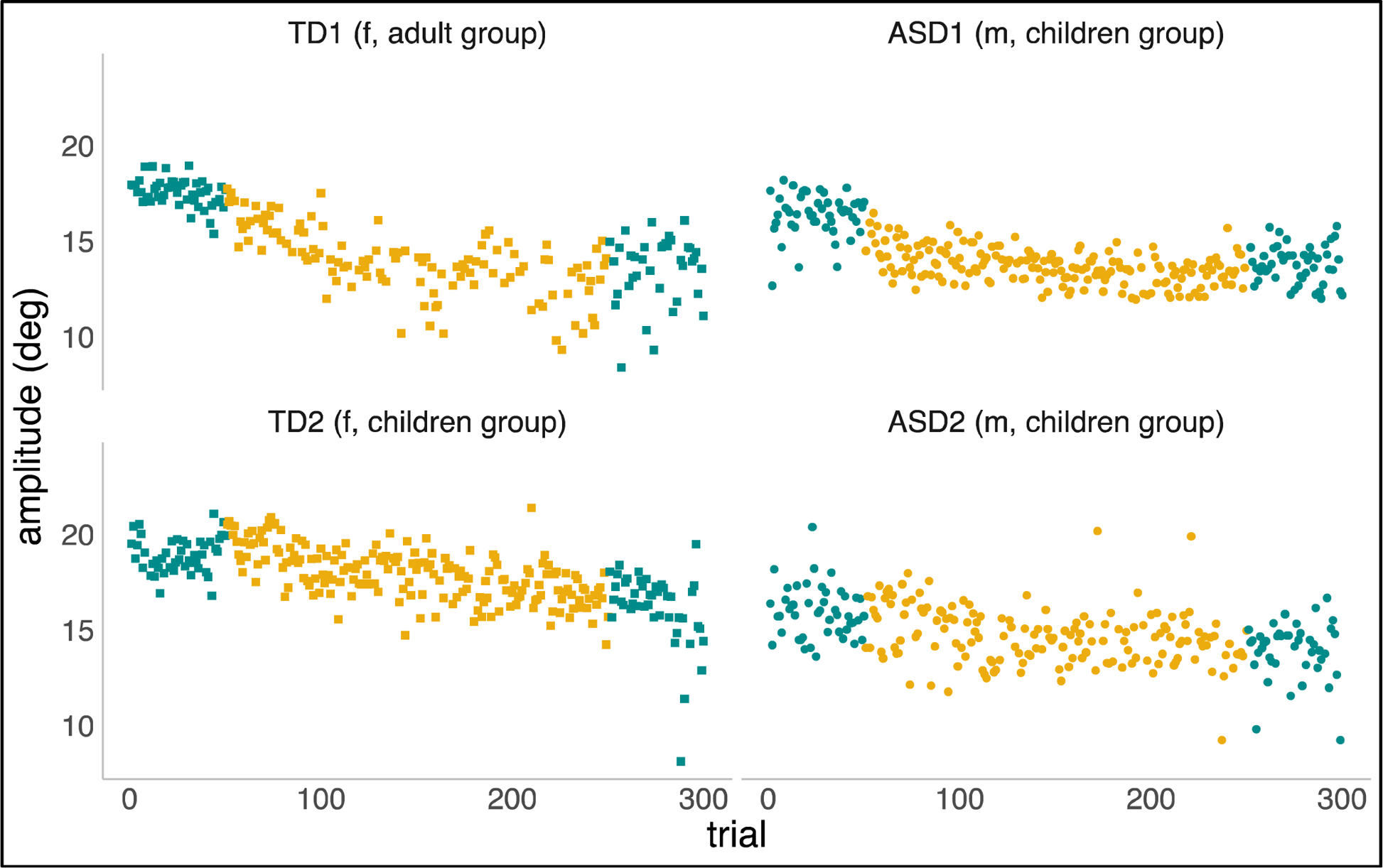
Saccadic adaptation examples to illustrate fast (top plots) and slow (bottom plots) adapters in both TD and ASD groups. The amplitude of primary saccadic eye movements is plotted as a function of trial number, i.e., the order in which they occurred during the experiment. Pre-adaptation or control trials (left) and post-adaptation or control trials (right) are plotted in blue. Adaptation trials are plotted in yellow.

We set out to examine the early phase (i.e., the response to the first 20 adaptation trials) versus the late phase (i.e., the last 20 adaptations trials) of adaptation, given prior work suggesting that adaptation might be slower to emerge in participants with ASD. Figure 2 (Children; 6-17 years old) and Figure 3 (Adults; 18-45 years old) show the mean amplitude of the baseline pre-adaptation trials, compared to the mean amplitude of the first twenty adaptation trials (Panel A1). In panel A2, the distributions of the differences between baseline and early-adaptation trials are plotted for each group (TD and ASD). The same plotting convention is used to show the comparison between baseline trials and the late adaptation trials in panel B of both figures.

**Figure 2.**
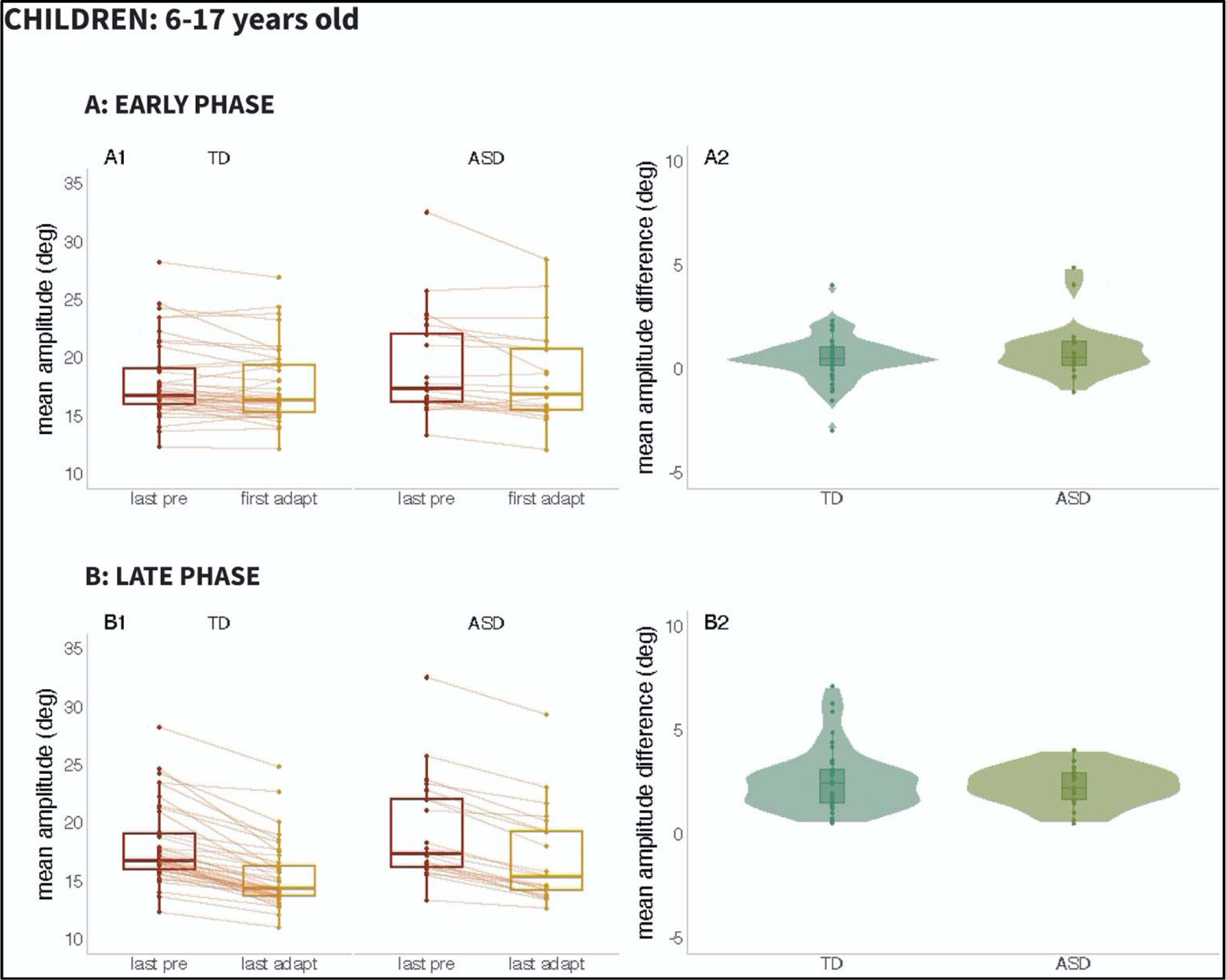
Results from Children (ages 6-17 years): <u>Panel A:</u> Early Phase for the 6-17 age range; (A1): Mean amplitude of the last twenty pre-adaptation trials and of the first twenty adaptation trials, (A2): Mean amplitude difference between pre-adaptation and adaptation trials from (A1). <u>Panel B:</u> Late Phase for the 6-17 age range; (B1): Mean amplitude of the last twenty pre-adaptation trials and of the last twenty adaptation trials, (B2): Mean amplitude difference between pre-adaptation and adaptation trials from (B1).

**Figure 3.**
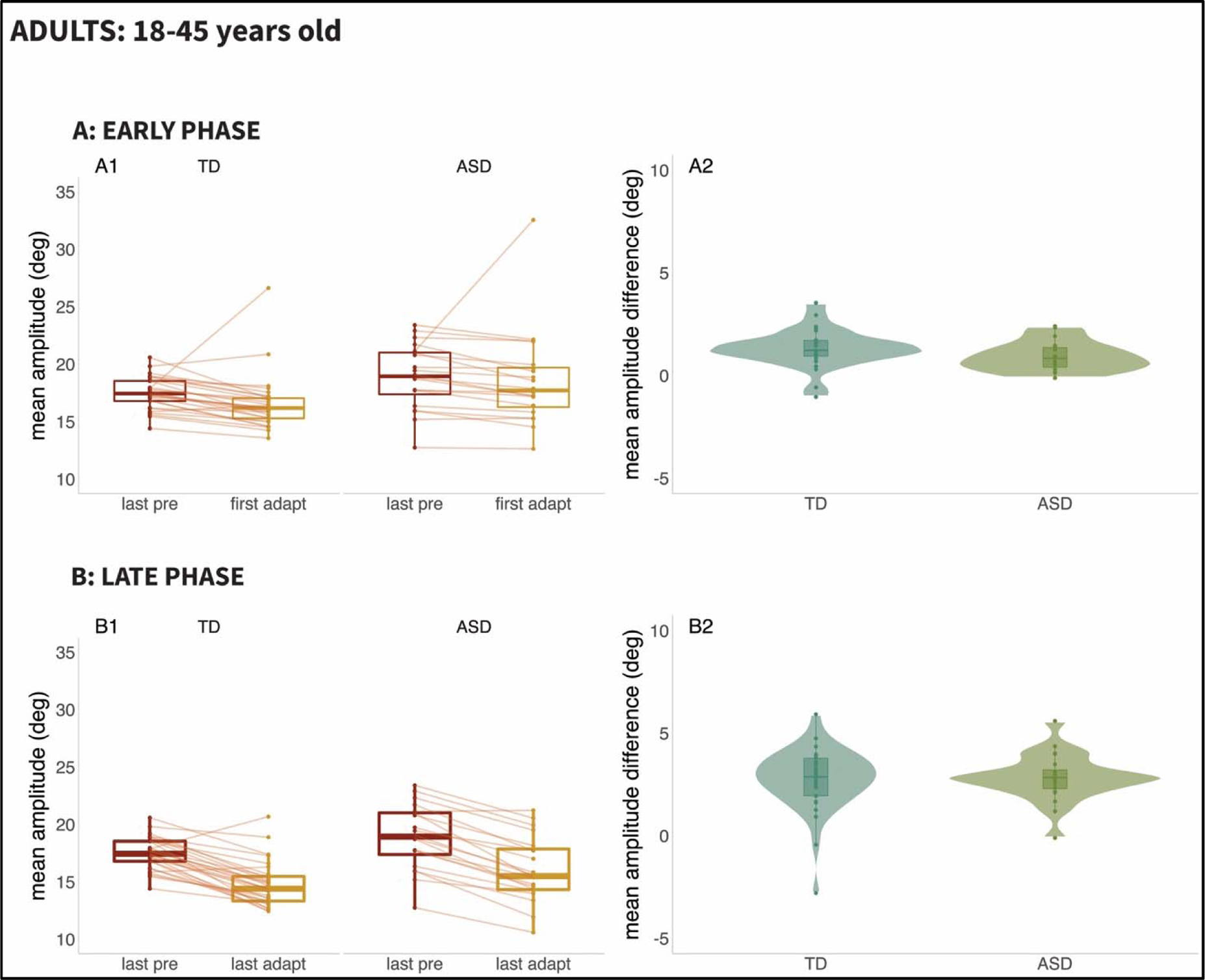
Results from Adults (ages 18-45 years): <u>Panel A:</u> Early Phase for the 18-45 age range; (A1): Mean amplitude of the last twenty pre-adaptation trials and of the first twenty adaptation trials, (A2): Mean amplitude difference between pre-adaptation and adaptation trials from (A1). <u>Panel B:</u> Late Phase for the 18-45 age range; (B1): Mean amplitude of the last twenty pre-adaptation trials and of the last twenty adaptation trials, (B2): Mean amplitude difference between pre-adaptation and adaptation trials from (B1).

A two-way repeated measures ANCOVA did not reveal effects of age (*F*(1,106) = 0.007, *p* = 0.936), of group (*F*(1,106) = 0.001, *p* = 0.981), or of the interaction between group and phase (*F*(1,106) = 0.401, *p* = 0.761) in saccade adaptation. However, the main effect of phase was significant (*F*(1,106) = 26.883, *p* < 0.001), reflecting the fact that the amplitude of adaptation was significantly higher in the late phase compared to the early one for both populations.

To test for differences in amplitude of eye movements between groups—that is, whether individuals with ASD tended to undershoot (hypometria) or overshoot (hypermetria) the target —we calculated the mean amplitude of all control trials in the opposite direction of adaptation trials and performed a univariate ANCOVA. Figure 4 shows individuals with ASD tended to make longer saccades to the target in this paradigm when compared to their age-matched controls. A statistically significant difference was found between ASD and TD groups in terms of amplitude of primary saccade when taking age into account: *F*(1,106)= 10.381, *p* = 0.002. The effect size of this difference was, nevertheless, small (η^2^= 0.089, TD: Mean=16.666; SD=0.351, and ASD: Mean=18.509; SD=0.452).

**Figure 4.**
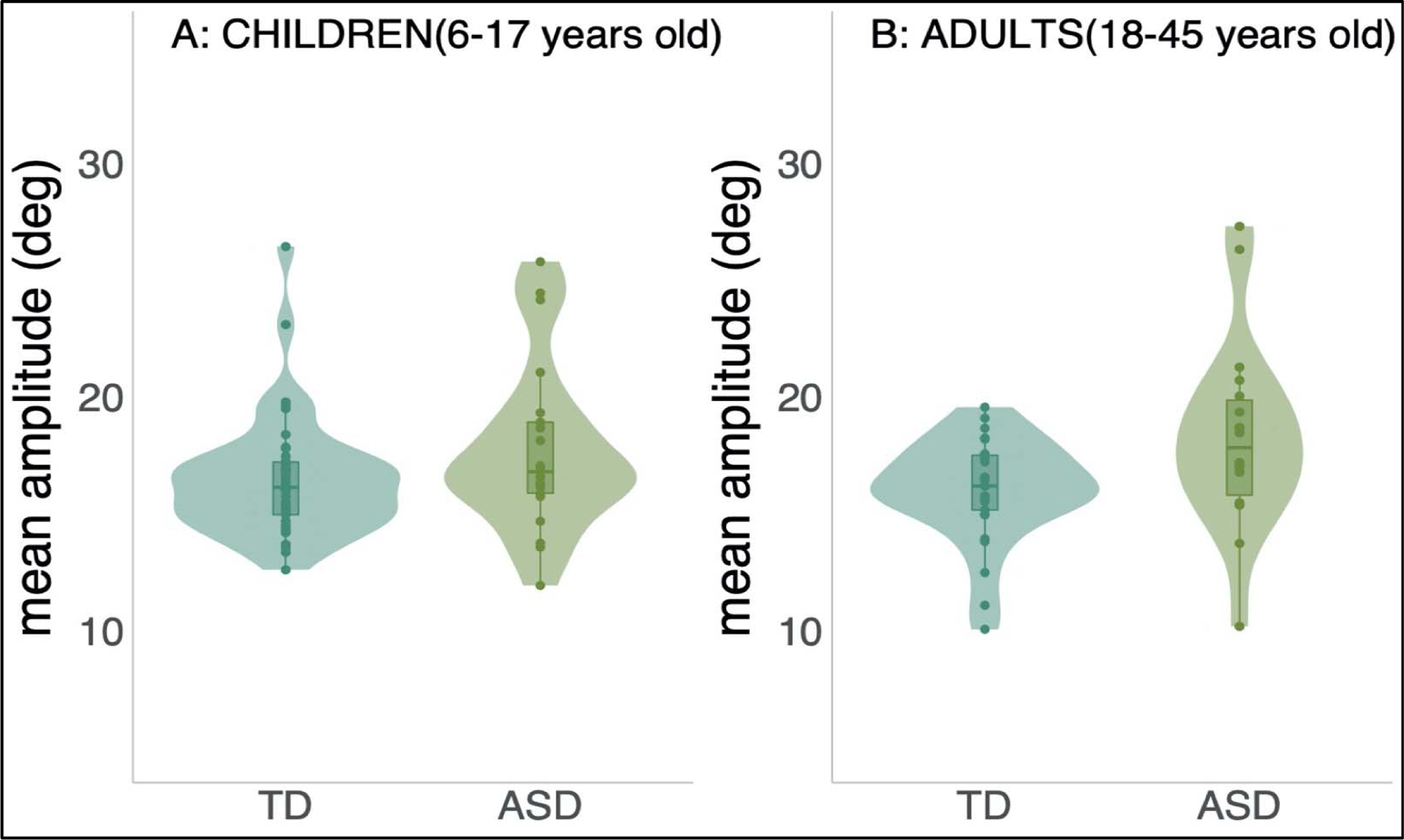
Mean amplitude of all control trials in the opposite direction of adaptation trials per group and both age range; (A): Children, 6-17 years old and (B): Adults, 18-45 years old.

Finally, an independent samples t-test revealed no statistically significant difference between the ASD and TD group in the normalized standard deviation based on the mean of the last 20 control trials for each individual participant (t(107)=-1.182, two-sided p= 0.240).

## DISCUSSION

The aim of this study was to investigate potential differences in saccade metrics between individuals with ASD and age-matched TD in the context of a classic gain-down saccadic adaptation paradigm (McLaughlin, 1967). The primary question was whether saccade adaptation to systematically introduced errors would be disrupted in this population and whether this might be more evident during the early phase of adaptation, as suggested by two previous studies in ASD populations (B. P. Johnson et al., 2013; M. W. Mosconi et al., 2013). We also measured the accuracy of saccades directed towards the hemifield opposite to that used for the adaptation task, to assess the presence of saccadic undershoot or overshoot in ASD. The primary analysis showed clear evidence for adaptation, both during the initial “early” phase (i.e., the first 20 adaptation trials), and again during the last 20 “late” adaptation trials. A main effect of phase confirmed that the later phase resulted in greater adaptation; that is, individuals continued to adapt across the adaptation trials. Our primary hypothesis, however, that sensory-oculomotor deficits would lead to slower and reduced saccade adaptation in ASD, was not supported by the data. This was found to be the case for both children and adults with ASD, providing no evidence for a developmental component. Also, in contrast to prior findings, there was no evidence that early adaptation was weaker in ASD, and again, this was the case for both the children and the adult groups.

In a secondary analysis, we assessed whether primary saccades directed towards the hemifield opposite to that used for the adaptation task would show anomalous saccade metrics. Here, we did find evidence for a tendency toward hypermetric saccades in ASD, but it should be emphasized that, while statistically significant, the effect size was found to be small. It should also be emphasized that we did not predict this finding and as such, it should be considered post-hoc and exploratory at this stage and would require replication before any strong conclusions should be reached. A finding of hypermetric saccades also stands in contrast to previous work, as the majority of studies that did find primary saccade differences, tended to report hypometria (N. Bast et al., 2021; B. Johnson et al., 2012; B. P. Johnson et al., 2013; Kovarski et al., 2019; Schmitt et al., 2014), although, there are also studies reporting no evidence for differences in saccade amplitude in ASD (B. P. Johnson et al., 2016; Kovarski et al., 2019; Luna et al., 2007; Minshew et al., 1999; Yukari Takarae et al., 2008).

Perhaps one of the more consistent findings in the literature on saccade metrics in ASD has been the finding of greater within-participant variability in saccade amplitudes (Glazebrook et al., 2009; Goldberg et al., 2002; B. Johnson et al., 2012; Minshew et al., 1999; Nowinski et al., 2005; Stanley-Cary et al., 2011; Y. Takarae et al., 2004). Here, in contrast, we found no evidence for this greater variability when we looked at the within-participant variance (normalized standard deviations) in primary saccades to the last 20 control trials that were delivered before the adaptation phase of the experiment was initiated.

What then of the putative link between cerebellar anomalies in ASD and eye-movement disturbances? One possible strategy would be to perform some stratification of the ASD population, either by first identifying a subgroup of individuals with ASD who show clear cerebellar structural differences on neuroimaging and asking whether this subgroup also show saccadic dysfunctions, or by separating out subgroups of ASD individuals who show anomalous saccade metrics (i.e., lack of adaptation, hypermetria) and asking if this subgroup would show greater instances of structural cerebellar differences. Of course, given the current results, instances of non-adaptation were just as common in the TD population as in the ASD, and the finding of a slight relative hypermetria in ASD stands in contrast to a number of previous studies that reported precisely the opposite.

Taken together, the current findings suggest that if there are differences in saccade metrics in ASD, these are of small effect size and very subtle. Considering our current findings and the lack of consistent evidence for saccade differences in the literature, it seems highly unlikely that there is a fundamental sensory-motor processing deficit in the oculomotor system in Autism. Of course, gaze and fixation differences have been widely observed and reported in Autism (Hirsch et al., 2022; Klin et al., 2002), leaving open the question as to what drives them. Although not tested here, it may well be the case that the alternate hypothesis, that these differences are related to social motivational factors, for instance a reluctance or difficulty in engaging with peers, is the more parsimonious explanation.

It is worthwhile considering what might account for differences in the results of the present study and that of Mosconi et al., the other relatively large sample study on saccade adaptation in ASD, where subtle saccade adaptation differences were reported for early stages of adaptation. Both studies had a considerable baseline period against which to assess saccade adaptation, used similar criteria for assessing early stages of adaptation (20 to 30 trials), and included ASD and TD participants of average and above cognitive ability over a similar age range. It is possible that differences in paradigm might be a factor. In the current study, the target was presented at 20 degrees from central fixation and reduced by 25% to induce saccade adaptation whereas in Mosconi et al., it was presented at 12 degrees and reduced by 16%. Thus, the use of targets of different eccentricity and the different magnitude discrepancies between T0 and T1 could contribute to the different results. However, in both cases, the vast majority of participants adapted, indicating that both were effective saccade adaptation paradigms. Whereas in our study, there was no significant group difference in number of individuals that did not adapt, in Mosconi, there was a substantial increase in the number of non-adapters in the ASD group and very few TD non-adapters. One possibility is that there is a trade-off between magnitude of T0 and T1 difference and initiation/speed of adaptation, and that larger magnitude differences are required, on average, for autistic individuals. Future work in which this parameter is varied will help to address this possible explanation. However, our ASD sample also did not show increased variability as it has been shown in some other studies, suggesting that differences between studies may reflect variability from sample to sample. The findings of the current study speak to a lack of robustness with regard to differences in ASD in saccadic adaptation, hyper-versus hypo-metric saccades, and saccadic variability, suggesting that, unfortunately, saccades and saccadic adaptation do not hold promise as early biomarkers for the detection of ASD.

## CONCLUSION

This study examined possible saccadic eye-movement differences between individuals with ASD and their age-matched TD peers in the context of a gain-down saccadic adaptation task. While studies reported in the literature have yielded mixed results regarding the ability of individuals with ASD to adapt saccade amplitudes in response to imposed visual errors, the data presented here provide no evidence for differential saccadic adaptation in Autism, either in children or adults on the spectrum. Nor was there evidence for a slower rate of adaptation in ASD in either children or adults. We also assessed whether primary saccades during initial control trials would be more variable at the individual participant level in ASD participants, but found no evidence for greater variability in this cohort. The data did point to significantly larger primary saccades to non-adapted targets in both children and adults with Autism, although this was a post-hoc finding and the effect size was small. Overall, in a sizable cohort of ASD children and adults (N=41), this study found scant evidence for differences in saccade metrics in Autism.

## Data Availability

Upon acceptance for publication, the authors will coordinate with the editorial office to ensure that the full and appropriately de-identified datasets are uploaded to a publicly available repository and that the appropriate link is made to the article file (e.g. Figshare, Dryad).

## Abbreviations

ASD: Autism Spectrum Disorder
TD: Typically Developing
ANCOVA: Analysis of Covariance
HFA: High Functioning Autism

## ACKNOWLEDGEMENTS

The authors acknowledge the contribution of University of Rochester undergraduate students Sri Nuvvula, Chloe Pacteau, and Suzan Hoffman, and of Albert Einstein College of Medicine study coordinator Elise Taverna for their assistance with recruitment and data collection, and of clinical psychologists Leona Oakes PhD and Juliana Bates, PhD for her contributions to phenotyping of participants.

## AUTHOR CONTRIBUTION

KT, JJF and EGF designed and implemented the study. KT, AAF and DH coordinated data collection. KT analyzed the data. AAF constructed the illustrations. SM took primary responsibility for coordinating recruitment and phenotyping of participants at the Albert Einstein collaborating site. KT wrote the first draft of the manuscript. All authors provided editorial input to KT on subsequent drafts. All authors read and approved the final version of this manuscript.

## DATA SHARING

Upon acceptance for publication, the authors will coordinate with the editorial office to ensure that the full and appropriately de-identified datasets are uploaded to a publicly available repository and that the appropriate link is made to the article file (e.g., Figshare, Dryad).

## COMPLIANCE WITH ETHICAL STANDARDS

The Research Subjects Review Boards of the University of Rochester and the Albert Einstein College of Medicine approved all the experimental procedures (respectively STUDY00002036 and 2009-523). All adult participants provided written informed consent and for all individuals below 18 their parents provided written informed consent and each participant in this age range provided written informed assent in accordance with the tenets laid out in the Declaration of Helsinki.

